# An MRI-based, data-driven model of cortical laminar connectivity

**DOI:** 10.1101/2020.06.02.20120303

**Authors:** Ittai Shamir, Yaniv Assaf

**Affiliations:** Department of Neurobiology, Faculty of Life Sciences, Tel Aviv University, Tel Aviv, Israel; Sagol School of Neuroscience, Tel Aviv University, Tel Aviv, Israel

**Author notes:** Correspondence: Ittai Shamir.

**Keywords:** cortical layers_1_, cortical connectivity_2_, magnetic resonance imaging_3_, cortical layer connectivity_4_, cortical modelling_5_

## Abstract

Over the past two centuries, great scientific efforts have been spent on deciphering the structure and function of the cerebral cortex using a wide variety of methods. Since the advent of MRI neuroimaging, significant progress has been made in imaging of global white matter connectivity (connectomics), followed by promising new studies regarding imaging of grey matter laminar compartments. Despite progress in both fields, there still lacks mesoscale information regarding cortical laminar connectivity that could potentially bridge the gap between the current resolution of connectomics and the relatively higher resolution of cortical laminar imaging. Here, we systematically review a sample of prominent published articles regarding cortical laminar connectivity, in order to offer a simplified data-driven model that integrates white and grey matter MRI datasets into a novel way of exploring whole-brain tissue-level connectivity. Although it has been widely accepted that the cortex is exceptionally organized and interconnected, studies on the subject display a variety of approaches towards its structural building blocks. Our model addresses three principal cortical building blocks: cortical layer definitions (laminar grouping), vertical connections (intraregional, within the cortical microcircuit and subcortex) and horizontal connections (interregional, including connections within and between the hemispheres). While cortical partitioning into layers is more widely accepted as common knowledge, certain aspects of others such as cortical columns or microcircuits are still being debated. This study offers a broad and simplified view of histological and microscopical knowledge in laminar research that is applicable to the limitations of MRI methodologies, primarily regarding specificity and resolution.

## 1 Introduction

In recent years, a growing number of studies have focused on techniques for exploring the brain’s wiring diagram, also known as the connectome (Hagmann 2009, Sporns 2012). Regarding global white matter connectivity (connectomics), diffusion weighted imaging (DWI) has become the standard imaging technique for exploring white matter tracts, by evaluating the directionality of water molecule diffusivity (Sporns et al. 2005, Paolo et al. 2007, Leemans et al. 2009, van Essen et al. 2013, Setsompop et al. 2013). DTI has certain inherent deficiencies and limitations in structural estimation, primarily regarding node delineation and edge mapping (Sotiropoulos and Zalesky 2019) and estimation of weaker or longer convoluted connections (Donahue et al. 2016), as well as a lack of directional or hierarchical information about interareal connectivity (Wang and Kennedy 2016). Nonetheless, diffusion MRI remains the primary in-vivo method for exploring whole-brain connectomics.

Significant progress has been made in the field of MRI neuroimaging of not only global white matter connectivity, but also cortical grey matter laminar structure. Regarding MRI estimation of grey matter, it has been established that myelination causes shortening of T1 values (Clark et al. 1992), consequently linking T1 to myeloarchitecture (Van Essen et al. 2019). Since then, studies have also pointed to correspondence between T1 clusters and cortical layers in rat brains (Barazany and Assaf 2012). Furthermore, it has been shown that low resolution T1 MRI can also be utilized to provide layer-specific information in human brains (Lifshits et al. 2018, Shamir et al. 2019). The integration of microstructural information concerning cortical properties and macrostructural connectomics could pose an exciting development in the field (Jbabdi and Johansen-Berg 2011, Johansen-Berg 2013).

In light of this progress, a generalization of a laminar model that can be integrated into connectomics has become essential. This model will hold the lacking mesoscale information regarding cortical laminar connectivity that could potentially bridge the resolution gap between macroscale connectomics and microscale cortical laminar imaging. This study offers a simple data-driven model of laminar connectivity that could be utilized to bridge this gap between two branches of neuroimaging and introduce a novel way of modelling whole-brain tissue level connectivity.

In order to form such a model of cortical laminar connectivity, we conducted a limited-scale systematic review centered around exploration of how cortical layers and their interconnections have been reported in peer-reviewed journals. It includes a systematic review of 51 peer-reviewed journal articles, including prominent studies and reviews centering around the subject of layer connectivity (see complete list of references), followed by quantitative and qualitative analysis of reported findings, including the variability and uniformity of each cortical partition. The results are offered as an unassuming straightforward model summary of reported findings, focused on cortical laminar connectivity on a whole system tissue level.

Study of the structure and function of the cerebral cortex started over two centuries ago using a wide variety of methods. Since then, great scientific efforts have been invested in uncovering the structural building blocks that comprise the cortex. These blocks include morphological and connectivity units, derived from horizontal and vertical partitioning of the cortex.

The cortex was first partitioned horizontally starting with the work of Gennari (Gennari 1782) and Baillarger (Baillarger 1840), leading up to the work of Korbinian Brodmann in 1909 (Garey 2006), which delineated six cortical layers distributed differently across cortical regions. Vertical (or radial) partitioning followed, with the introduction of cortical columns as units that span all cortical layers in the 1950s (Lorente de No 1949, Mountcastle 1957) and by Hubel and Wiesel throughout the 1960s (Hubel and Wiesel 1959, 1962, 1965, 1968, 1969). The cortical column came as a generalization of Hubel and Wiesel’s discovery of ocular dominance columns in the mammalian visual cortex. Last to be introduced were the cortical connectivity units across horizontal and vertical components, also known as the cortical microcircuit (canonical microcircuit). The concept of the microcircuit first appeared in the 1970s, following the work of Szentagothai (Szentagothai 1975), Creutzfeldt (Hellweg et al. 1977) and Wiesel (Gilbert and Wiesel 1983). Throughout the 1980s, a broader point of view was offered regarding cortical layer connectivity of the macaque visual cortex (Rockland and Pandya 1979, Maunsell and Van Essen 1983, Felleman and Van Essen 1991). As a rule of thumb, the canonical microcircuit comprises of thalamic input to granular layer 4, which projects to superficial layers 2 and 3, which in turn project to deep layer 5 and then layer 6, which projects back to the thalamus (Dhruv 2015).

While these three partitions of the cortex into horizontal, vertical and connectivity units appear to be logical extensions of one another and are often referred to as common knowledge, it appears that certain aspects of some have yet to be “set in stone”. While it has been widely accepted that the cortex has an exceptionally organized and interconnected structure, published articles on the subject display a variety of approaches towards its structural building blocks. Von Economo even notably stated that: “the distinction of six layers can be both arbitrary and conventional. Nevertheless, on practical grounds, we retain the six-layer division” (Von Economo 2009). When addressing the number of cortical layers in a column, there lacks a uniformity in grouping of the layers. To this day, studies discuss different groups of cortical layers (Bosman and Aboitiz 2015, Roelfsema and Holtmaat 2018) and the concepts of the cortical column and microcircuit have long been debated (Nelson 2002, Dhruv 2015), some even offering alternatives such as micro- and macro-columns (Mountcastle 1957, Buxhoeveden and Casanova 2002), or modifications such as region-specific microcircuits (Shipp 2007, Shepherd 2011, Raizada and Grossberg 2003). In other words, it appears that there lacks a consensus around a single unified layer model.

The lack of a broad and cohesive viewpoint of cortical units in general, and cortical microcircuits in particular, can be attributed to the wide variety of methods which are typically used in a region-specific outlook. This variety of methods starts with the diversity of animal models used, including but not limited to cats (Hubel and Wiesel 1962, Douglas and Martin 1989) and non-human primates, such as macaque and rhesus (Felleman and Van Essen 1991, Mountcastle 1997). Other animal models include rodents, mainly rat and mouse but also rabbit and opossum, and sauropsids (birds, reptiles, turtles, etc.). Not only that, but many published reviews also combine data from multiple species in a single model. The methods used for assessing the cortical units in the animal model are even more broadly varied, including histological methods (Golgi, Nissl), different types of retrograde and anterograde tracers, electrophysiological recording, microscopy (ESEM, IR microscopy), photonics, imaging, simulations and many more. In order to present a clear and unified model of laminar connectivity that fits our needs, we offer a simplified data-driven model that integrates white matter and grey matter MRI datasets into a novel way of exploring whole-brain laminar-level connectivity.

We hypothesize that MRI, or more specifically diffusion weighted imaging (DWI) and inversion recovery echo planar imaging (IR EPI), can be combined with histological findings regarding cortical layer connectivity, in order to model networks of cortical laminar connectivity.

## 2 Methods

Other reviews and integrative studies of cortical connectivity exist, offering a significantly wider viewpoint of the cognitive architecture underlying non-primary cortical regions (Solari and Stoner 2011) or multi-scale connectivity of vision-related cortical regions (Schmidt et al. 2018) in the primate brain. These studies offer a deeper dive into integration of findings regarding axonal tracing and cortical architecture through histological means. We chose to conduct a limited-scale review of 51 articles, with the distinct and novel purpose of forming a simplified whole-brain model that is directly applicable to the limitations of MRI datasets. Our data-driven model is based on a systematic review of publications, including prominent studies and reviews, centering around the subject of cortical laminar connectivity (see complete list of references). Articles were chosen according to relevance to the topic of cortical layer connectivity, preferably choosing those that include a schematic model of the rules of connectivity (see figure 1).

**Fig. 1.**
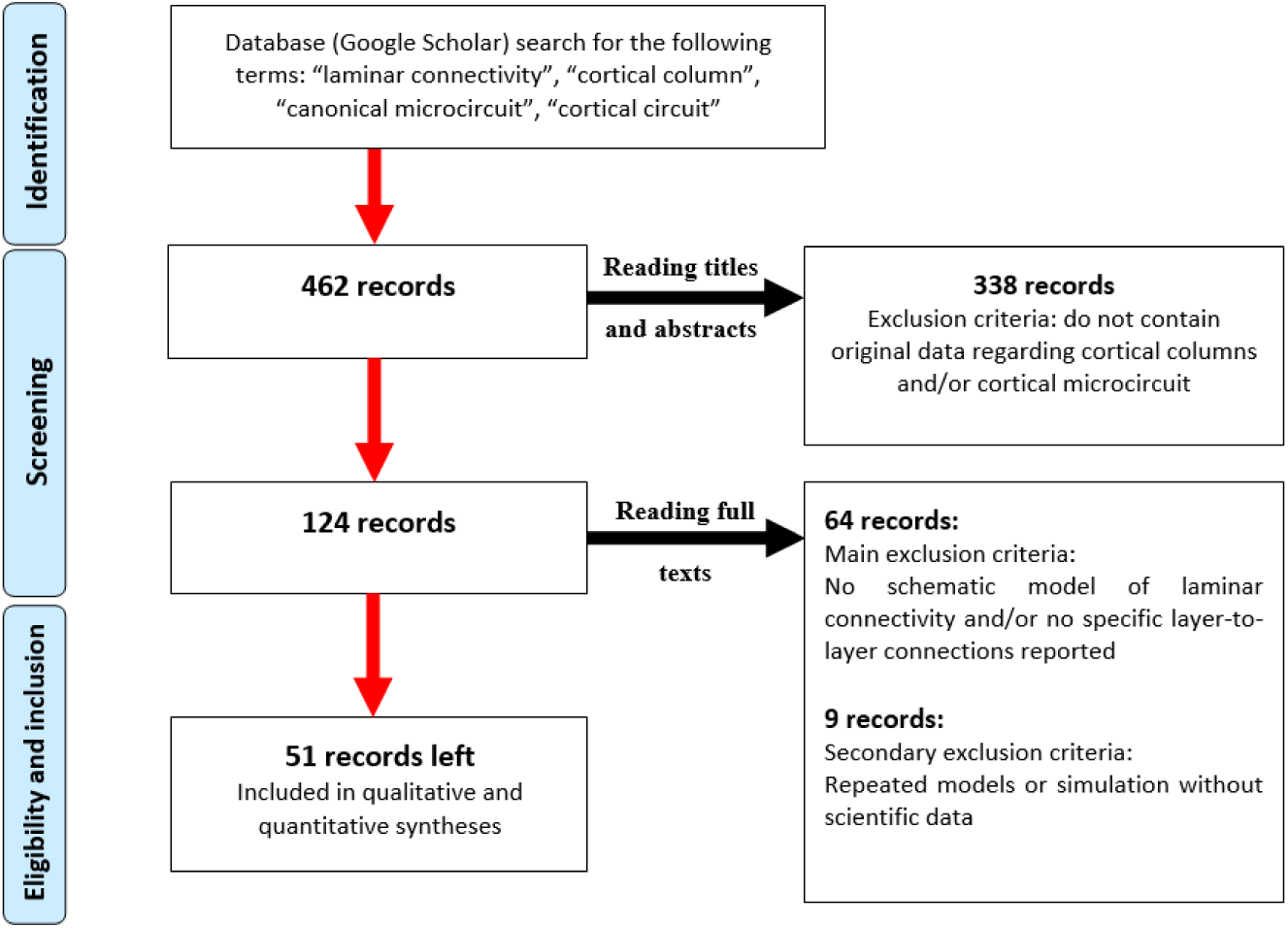
Flow chart of the selection of records (adapted from Moher et al. 2009)

The 51 reviewed articles were published between 1962 and 2018, but the overwhelming majority was published since 2000. Each article was methodically examined for the following criteria (for a summary of all included articles see Electronic Supplementary Material, section 1, table 1):

1. Reference data: authors, year, title, journal, volume, pages, DOI
2. Human/animal: subjects used for model
3. Method: methods used for visualization and modelling
4. Model (yes/no): schematic model included
5. Region definition: cortical regions discussed
6. Layer definition: definition of the cortical layers
7. Connection types: reported types of connections
8. Canonical circuit (pro/against): general tone concerning canonical circuit
9. Connection origin and termination: region and layer origin and termination (repeated for each reported connection)

In order to conduct a quantitative analysis of the reported cortical laminar connectivity, we first had to standardize the data between the articles included. First and foremost, we standardized data entry by examining both written connections and schematic model connections while making sure not to repeat identical connections in a single article. Secondly, we standardized data analysis by converting all cortical layer reporting of origins and terminations of connections to Brodmann’s six-layer definition.

Once the data standardization was completed, we conducted quantitative and qualitative analyses of each criterion and chose a simplified modelling framework. Our framework includes a set of whole-brain laminar-level connectivity rules that integrate macroscale connectomics with mesoscale cortical composition. The rules are applied mainly on DWI white matter tractography, according to the locations and granularity indices of the connecting regions and weighted according to their respective cortical laminar compositions. A smaller portion of the rules are applied on assumed connections, which are not visualized beyond the resolution of tractography. Overall, our model is composed of the following cortical building blocks:

1. Cortical layer definitions: grouping of the layers according to reported definitions.
2. Vertically oriented connections: extrinsic vertical connections between cortical layers and the subcortex (tractography-based) and intrinsic regional connections between the layers (an adaptation of assumed connections in the canonical circuit).
3. Horizontally oriented connections: extrinsic horizontal connections between cortical layers, including both intra- and interhemispheric connections (tractography-based).

Our final model is then implemented on the following input datasets:

1. White matter connectivity-standard diffusion weighted imaging (DWI) dataset, analyzed for tractography using constrained spherical deconvolution (CSD) in ExploreDTI (Leemans et al. 2009).
2. Grey matter laminar structure- published histological dataset on cortical layer composition across the whole brain (based on Scholtens et al. 2016).

## 3 Results

This section will open with a full description of the components of our model of cortical laminar connectivity, chosen based on the corresponding results of our systematic review, and close with an exemplary implementation of the model, expanding a standard connectome to a laminar connectome.

### 3.1 Model components

After conducting our systematic review of a selection of articles, the following cortical building blocks were chosen:

#### 3.1.1 Cortical layer definitions

When addressing horizontal partitioning of the cortex in a column or circuit, several varying layer definitions exist, sometimes appearing within a single article or review. In order to examine layer reporting throughout our systematic review, we used the six-layer definition. Overall, L2 and L3 are most often batched together as L2/3, followed by L5 and L6 batching as L5/6. Regarding subdivision of layers into sublayers, L1 and L2 are the least likely layers to be further divided. L3 and L5 are most often subdivided into two to three sublayers each, followed by L4 and L6 that are also occasionally subdivided into two to three sublayers. L4 is infrequently further subdivided into up to four layers, principally in the primary visual cortex. Consequently, grouping of the layers also varies from rougher grouping into three groups, including supragranular (L1-L3), granular L4 and infragranular (L5-L6), up to finer grouping into up to eight or more groups. Other grouping variations exist, including everything in between, but typically naming five to seven layers.

On the topic of cortical laminar components, our model uses the rough grouping of layers based on granularity, into infragranular, granular and supragranular layers. The granularity-based categorization appears to be the most robust across our sample of selected articles. It is worth noting that granularity-based grouping of the layers has also been linked to functional significance, suggesting that the granularity-based components exhibit not only anatomical interconnectedness, but also functionally distinct processing (Barone et al. 2000, Bastos et al. 2012). The general approach towards each of the six cortical layers from the “classic” Brodmann definition can be summarized in table 1 (below).

**Table 1.**
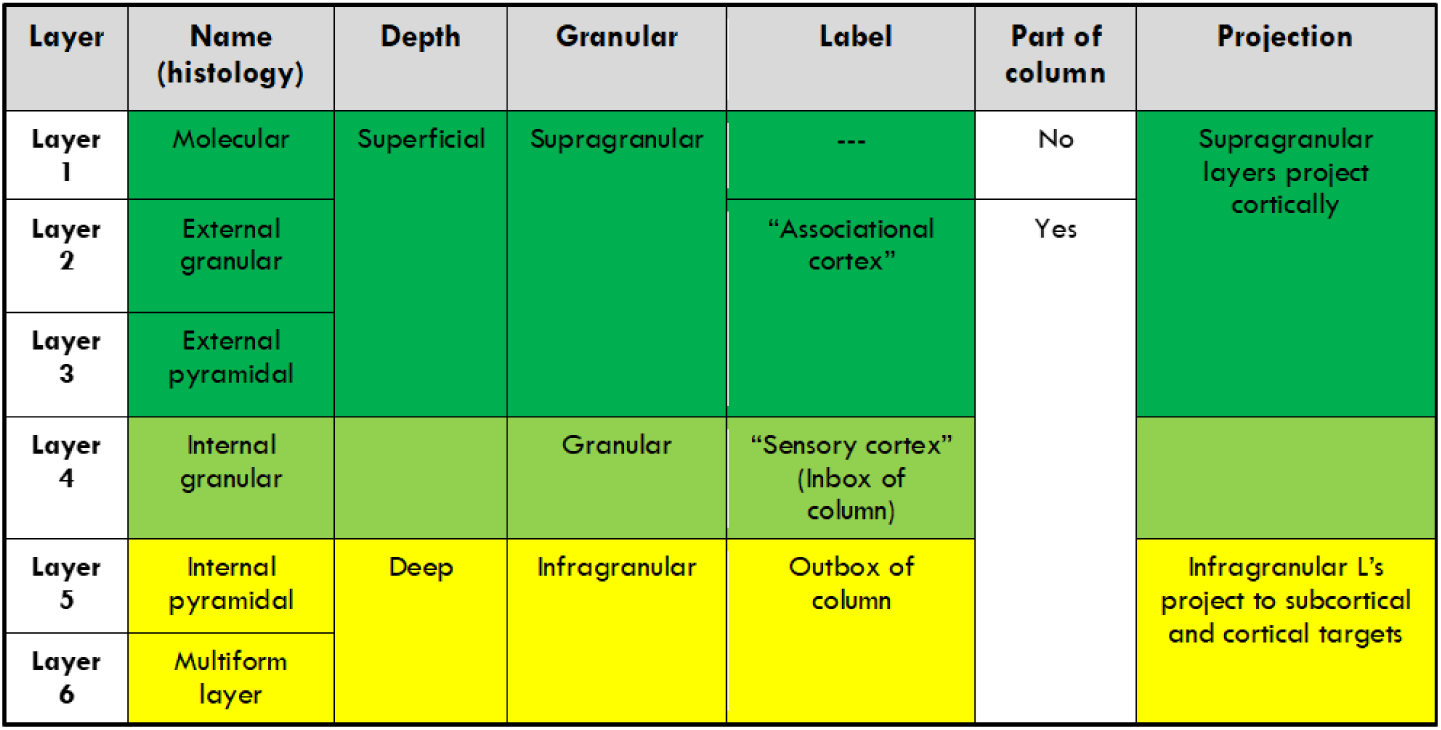
General approach towards the “classic” six cortical layers according to different criteria

#### 3.1.2 Vertically oriented connections

Vertically oriented connections include single-hemisphere intraregional connections between cortical layers and subcortical regions. The canonical microcircuit is the basic model of these layer-to-layer connections, and it is innately related to the concept of the cortical columns. Accordingly, an inherent bias would be expected in our chosen articles towards both the cortical column and the canonical microcircuit. Nevertheless, our analysis demonstrates a mixed approach towards both concepts. The relation of cortical columns to connectivity modelling has been put in doubt, with some commentaries going as far as writing ‘obituaries’ for the column and calling it ‘a structure without function’ (Feldmeyer 2012).

This review validates the ambivalence expressed towards cortical columns and microcircuits (for a summary of the approach of the review articles towards the canonical circuit see figure 2). Articles regarded as ‘pro’ mention the circuit by name and present a generally positive approach towards it or a modified version of it. For example, (Shipp 2007) suggests that although some details may still be unclear, it is reasonable to assume that a basic unit such as the canonical circuit exists repeatedly throughout the brain, creating exemplars like face selective neurons. Articles regarded as ambivalent also mention the circuit by name but present a more nuanced and critical approach towards it. For example, (Dhruv 2015) points out that while the canonical model exists with good reason, new departures from this standard keep popping up in publications. Articles regarded as against the canonical circuit mention it by name and present a directly critical approach towards it. For example, (Guy and Staiger 2017) claim that the subject of the layers is riddled with ambiguity, resulting in the interchangeable use of the terms ‘layers’ and ‘circuit’ and thus inadvertently assigning a function carried out by a circuit to a specific layer. Articles categorized as ‘no mention’ avoid use of the term and focus on other issues, such as extrinsic circuitry (Felleman and Van Essen 1991) or development and cell migration (Kennedy and Dehay 2012).

**Fig. 2.**
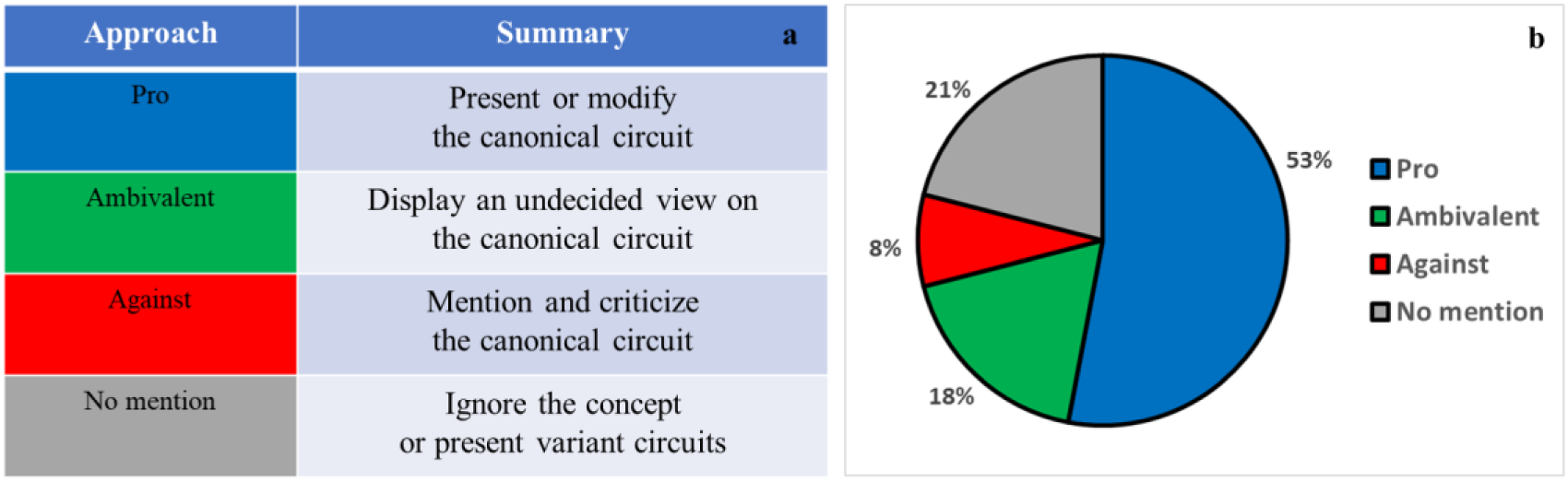
Approach of reviewed articles towards the concept of the canonical circuit: detailed table summary (a) and by percentage (b)

In the context of cortical laminar connectivity, it appears the subjects of both cortical regions as well as cortical layers are most commonly addressed through the lens of granularity. The granular visual cortex was the most discussed cortical region, followed by a general cortical model and a somatosensory model (see figure 3a). With regards to cortical layers, mid-cortex layers were the more commonly discussed layers, compared to agranular layers such as L1 (see figure 3b). Consequently, our model uses a granularity-based cortical atlas. Because of the lack of region-specific reporting on cortical laminar connection, a generalized granularity-based atlas such as the von Economo- Koskinas atlas best fits our model. This labelled atlas divides the cortex into regions based on their granularity index, ranging from agranular low order cortex to high order granular cortex (see figure 3c).

**Fig. 3.**
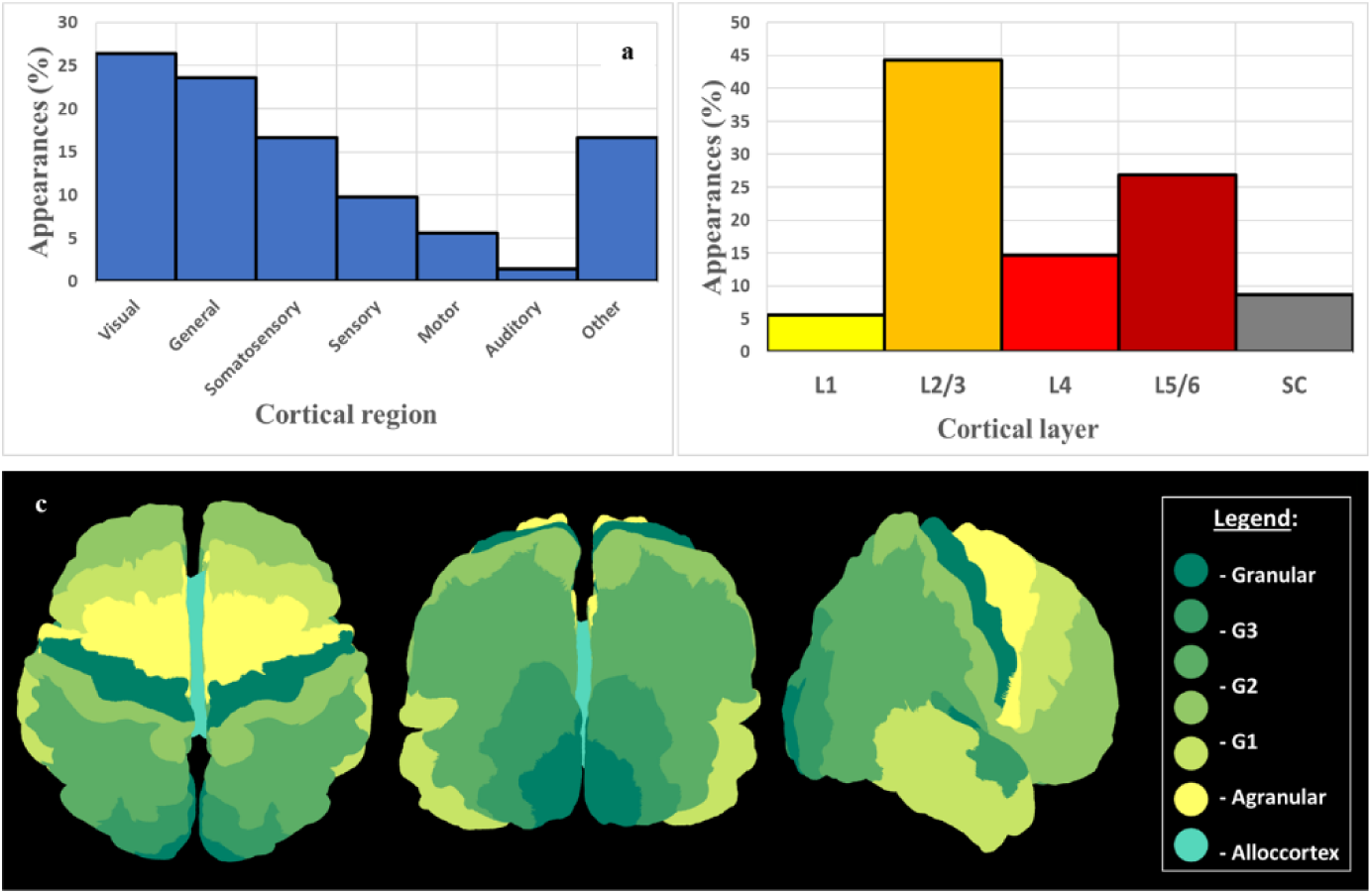
Cortical region (a) and cortical layer (b) reporting in laminar connectivity context in the articles reviewed in our systematic review (SC- subcortex), followed by a cortical atlas (c) labelled according to granularity index (similar to an adapted von Economo- Koskinas atlas shown in both Solari and Stoner 2011 and Beul and Hilgetag 2015)

In order to build a data-driven adaptation of the canonical microcircuit, we examined the distribution of all reported cortical laminar connections across all regions (see figure 4, for summary of findings for other regions see Electronic Supplementary Material, section 2, figures 1–5). This summary of reported laminar connections is organized by origin and termination of connections, using the three-layer grouping of the cortical layers into infragranular, granular and supragranular layers. Overall, this pattern of laminar connections follows the canonical circuit rule of connectivity:

1. High probability of connections that originate in subcortical regions and terminate in the granular layer (figure 5a).
2. High probability of connections that originate in the granular layer and terminate in supragranular layers (figure 5b).
3. High probability of connections that originate in supragranular layers and terminate in supragranular layers (figure 5c).
4. High probability of connections that originate in supragranular layers and terminate in infragranular layers (figure 5d).

**Fig. 4.**
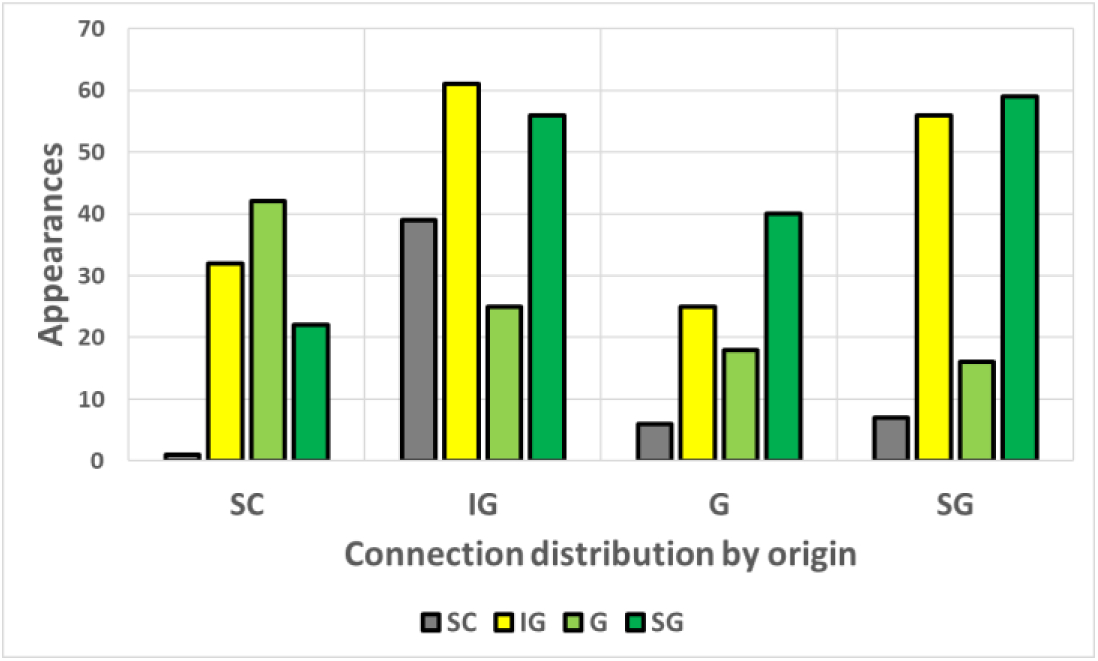
Summary of all reported cortical laminar connections, presented as distributions of connection terminations that originate in each of the following: subcortex (left), infragranular (mid left), granular (mid right), supragranular (right)

The vertically oriented cortical connections incorporated in our model are summarized below in an adaptation of the canonical microcircuit (see figure 5). The extrinsic connections, including cortex to subcortex connections, are based on DWI tractography and assumptions are made to the laminar level (component a in figure 5). The intrinsic connections on the other hand, including connections between two local laminar components, are beyond the resolution of tractography and are therefore assumed using weighted laminar components in the cortical region (components b-d in figure 5). In other words, when applying our model of cortical laminar connectivity, connection strengths are weighted according to corresponding laminar components.

**Fig. 5.**
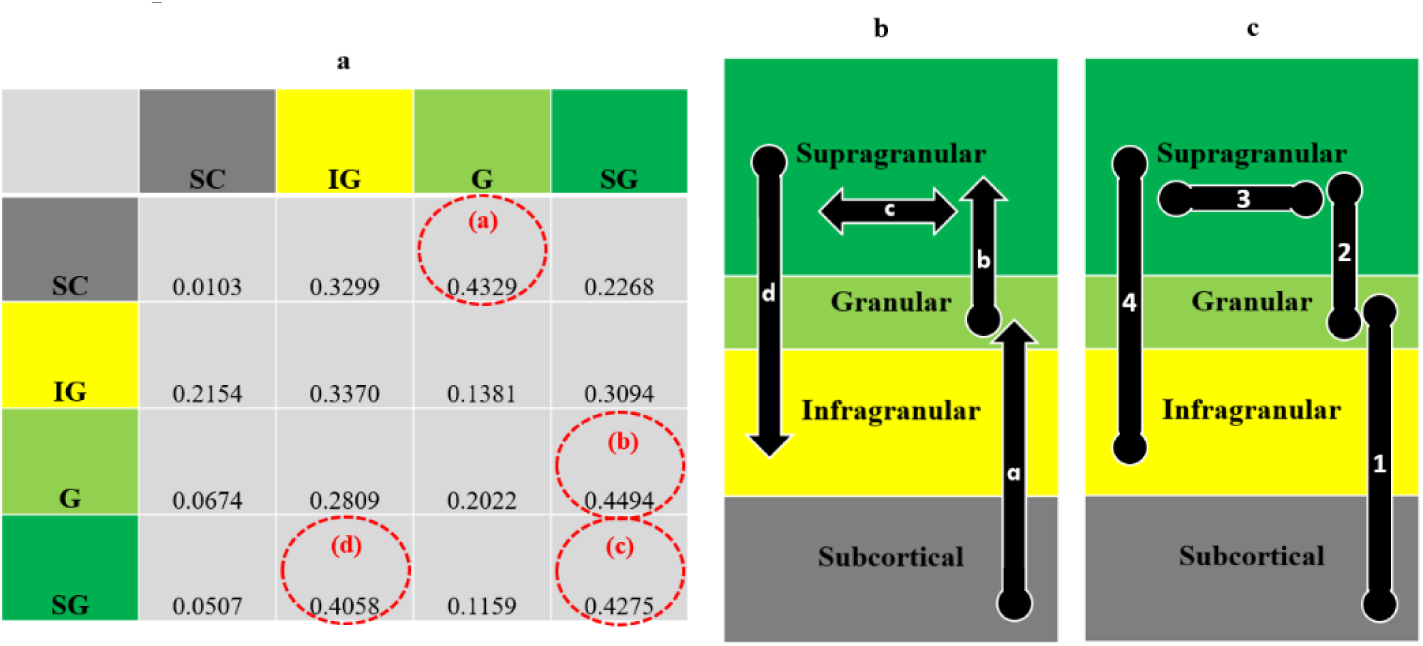
A summary of all reported vertically oriented cortical connections: a- Connectivity matrix, where: [row, column] = [connection origin, connection termination] and connections are averaged per origin (per row) b- A schematic summary of the most highly reported cortical laminar connections between granularity-based laminar groups, where: a-d connections correspond to a-d components in matrix c- A simplified undirected summary of (b)

#### 3.1.3 Horizontally oriented connections

Horizontally oriented connections are extrinsic nonlocal connections between laminar components of different cortical regions. Rules for horizontal connections in our model were extracted and expanded from a granularity-based model, which has been repeatedly reported and studied since the 1980s (Beul and Hilgetatg 2015, adapted from von Economo 2009 and from Barbas and Rempel-Clower 1997; a similar model also appears in Shipp 2005, adapted from Barbas and Rempel-Clower 1997 and Barabs 1986). The reported model delineates laminar origins and terminations of horizontal connections according to the granularity index of the corresponding cortical regions. Our model uses the granularity indices of the connecting regions (as shown in figure 3c), in order to apply the fitting rule of horizontal connectivity. As with the vertical connections (section 3.1.2), connection strengths are weighted according to corresponding laminar components (for an expanded explanation of the origin of our rules for horizontal laminar connections, see Electronic Supplementary Material, section 3).

Horizontal connections include the following two categories:

1. Intrahemispheric connections- between laminar components of a single hemisphere: The rules of connectivity were modeled using layer and region components according to the granularity indices of the connecting cortical regions (see table 2a, 2b). Our model of cortical laminar connectivity is a simplified undirected version of the aforementioned well-studied model (for the full set of rules see table 2c).
2. Interhemispheric connections- between laminar components of opposite hemispheres: Interhemispheric connections both originate and terminate in layers according to their corresponding granularity indices layers (Lodato and Arlotta 2015, Hawkins et al. 2017). Accordingly, the appropriate rule of connectivity could be considered a specific case of the abovementioned table (rule c in table 2b).

**Table 2.**
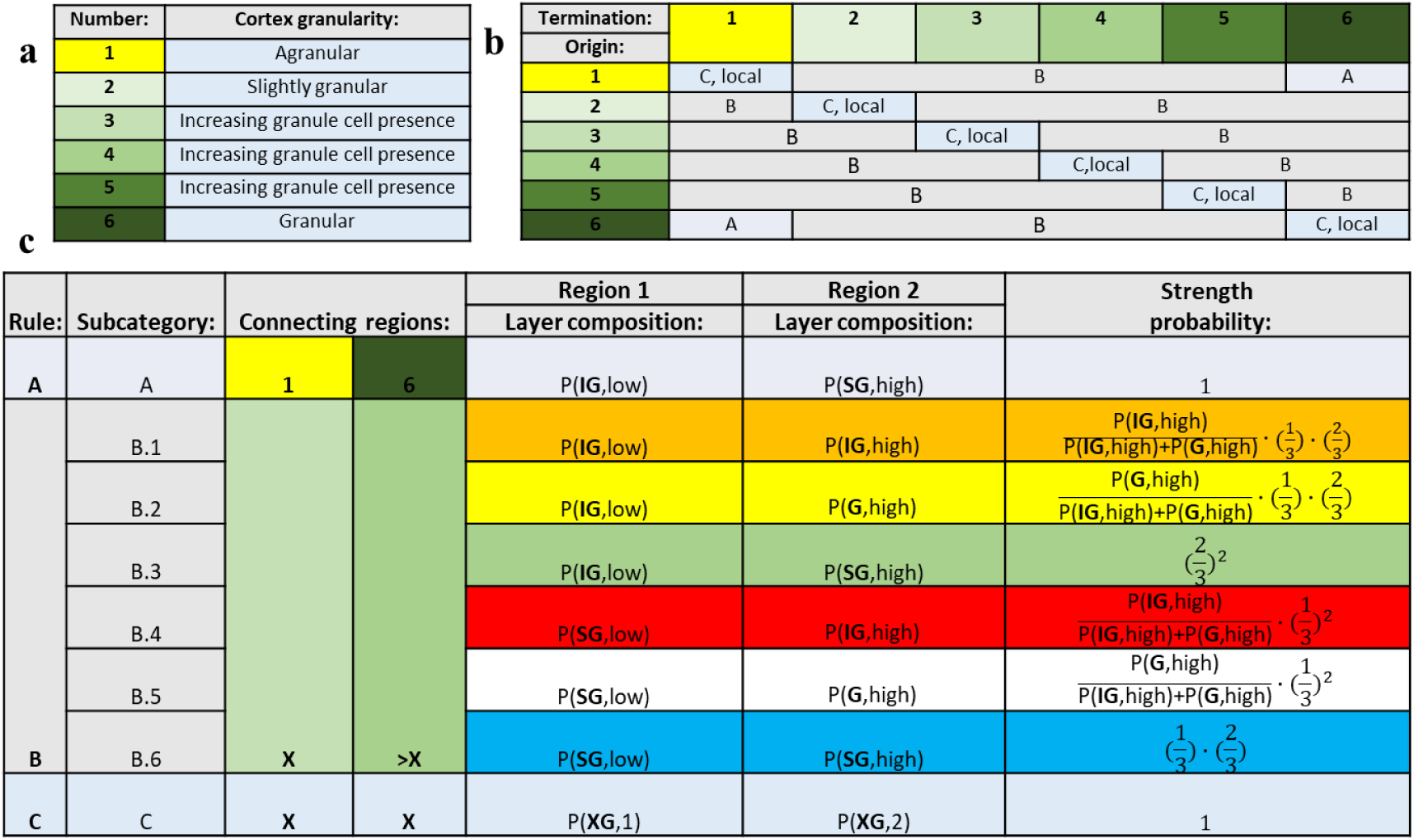
Our model’s rules of horizontally oriented laminar connections: a- Granularity indices of the connecting cortical regions: ranging from agranular (1) to granular (6) b- Rules of undirected laminar connectivity (A-C): categorized according to the granularity of the connecting regions c- Full description of all rules of undirected connectivity: estimated according to their laminar compositions Final laminar connection strengths are calculated by multiplying tractography connection strengths by laminar strength probabilities (last column) Where: P(L,R)- composition of laminar group L (SG- supragranular, G- granular, IG- infragranular) in region R (high/low represents relative granularity index, or 1/2 when they are equal)

### 3.2 Model application

Our model’s input datasets include information regarding both cortical connectivity as well as laminar structure:

1. Global white matter connectivity- standard DWI tractography and connectivity analysis using von Economo Koskinas (VEK) atlas (Scholtens et al. 2016, see figure 6a, 6b).
2. Grey matter laminar structure- supragranular, granular and infragranular laminar components across all von Economo Koskinas atlas regions (see figure 6c). As previously stated, great strides have been made in MRI estimation of cortical grey matter laminar structure (Barazany and Assaf 2012, Lifshits et al. 2018, Shamir et al. 2019). Nonetheless, we chose to use histological data that has already been published (based on Scholtens et al. 2016) in order to simplify the presentation and application of our model.

**Fig. 6.**
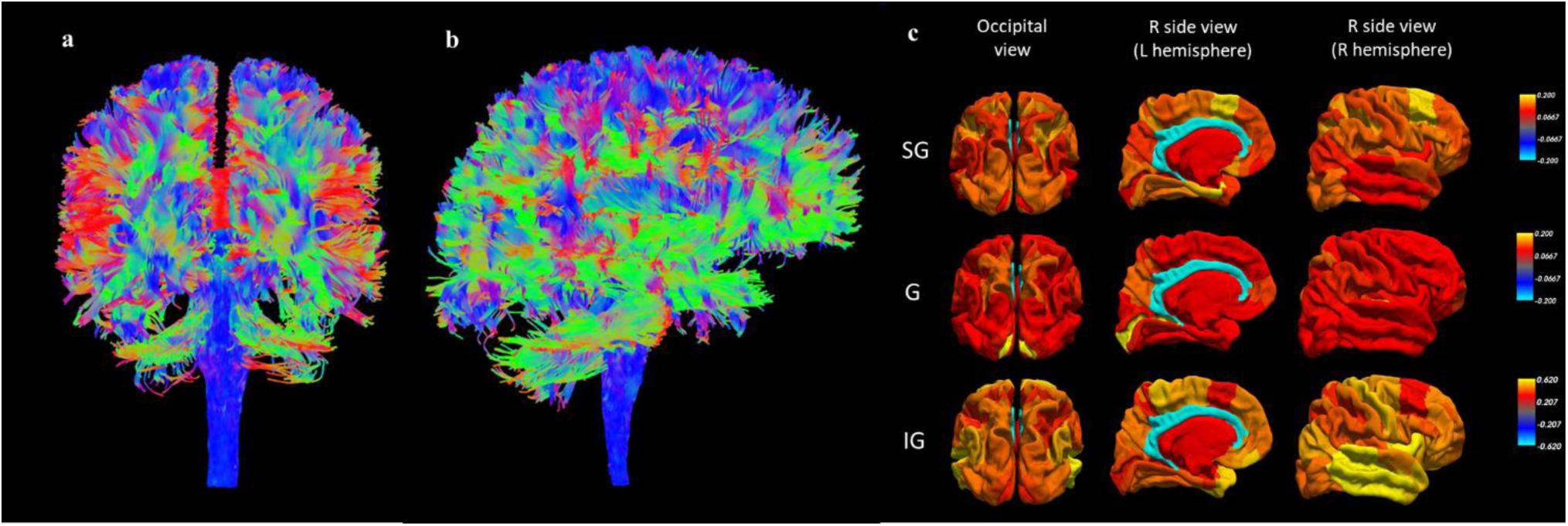
Model input datasets: Global white matter connectivity (visualized using ExploreDTI, Leemans et al. 2009): a- Tractography occipital view b- Tractography right side view c- Grey matter laminar structure (based on 6-layer published data from Scholtens et al. 2016): A summary of cortical laminar components across von Economo- Koskinas regions: supragranular (SG), granular (G) and infragranular (IG) layers

The cortical granularity indices (as shown in figure 3c) and the white and grey matter MRI datasets were all analyzed in the VEK atlas space (for a comprehensive network representation of all our model’s input datasets see figure 7 a-d). All white matter connections were then clustered using a standard k-means algorithm, based on a vector of the following properties for each connection: connection length, connection strength and the cortical laminar composition of the connecting regions. The process resulted in four robust clusters (see figure 7 e), displaying a visual correspondence to the following three fiber categories: cluster 1 corresponds to interhemispheric connections (commissural fibers), clusters 2–3 correspond to connections between the cortex and the subcortex, and cluster 4 corresponds to intrahemispheric connections (association fibers).

**Fig. 7.**
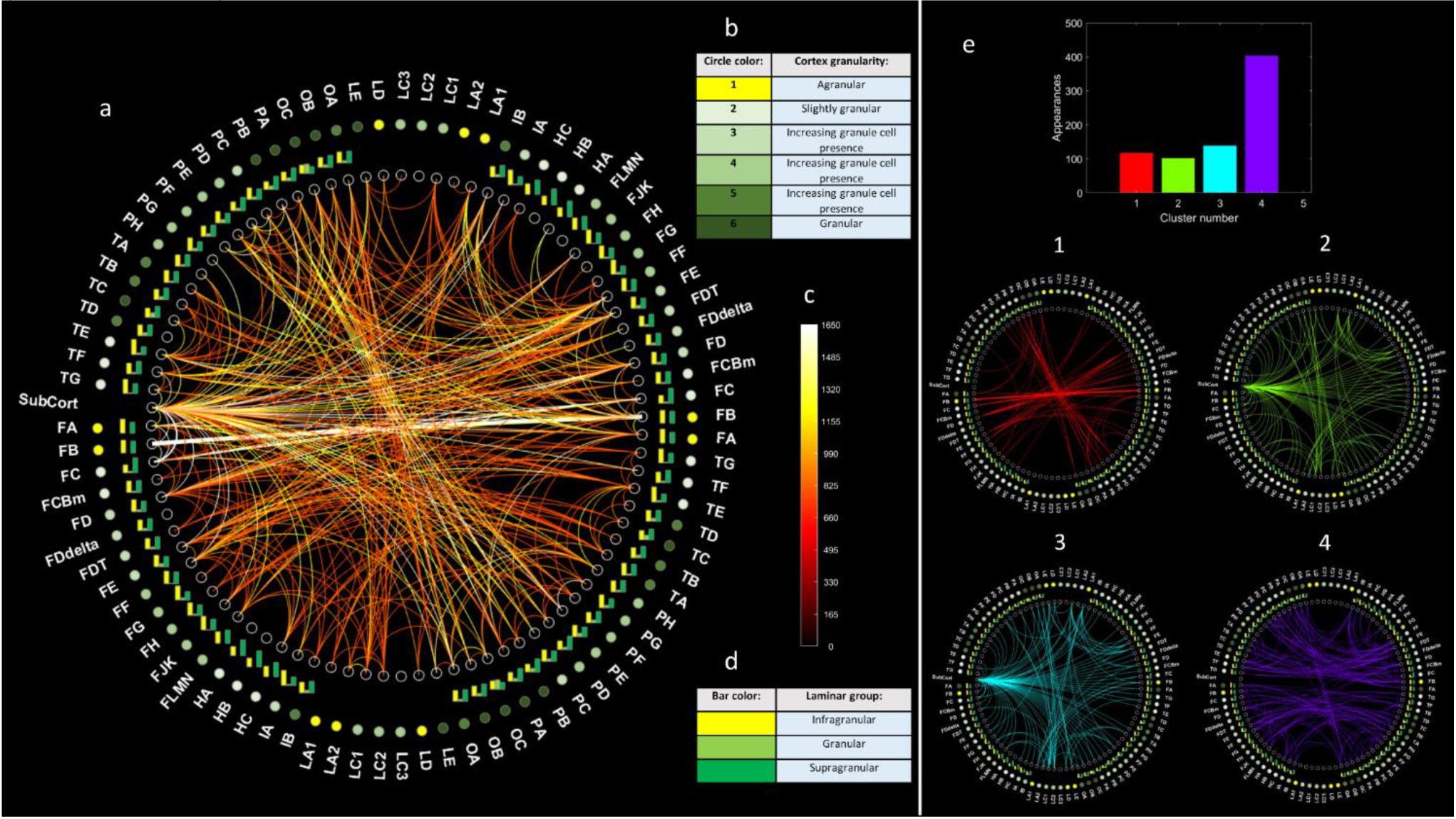
A circular network graph of all model input datasets (a), including: a- Legend for granularity indices, located in the outermost circle; b- Color scale for global white matter connectomics, representing connection strength of edges; c- Legend for color bars, representing grey matter laminar composition, located in the first outer circle; d- K-means clustering of global white matter connectomics, where edges are clustered using k-means based on the connection length and strength and the laminar composition of the connecting regions: cluster histogram (top) and each of the 4 individual clusters (1–4) (visualization completed based on: Paul Kassebaum (2020), circularGraph (https://www.github.com/paul-kassebaum-mathworks/circularGraph), GitHub)

Using the abovementioned white and grey matter datasets in the von Economo- Koskinas atlas space, we applied our model of cortical laminar connectivity using the following steps:

1. Tractography connections: vertically oriented connections between the cortex and the subcortex, and all horizontally oriented connections were expanded to the laminar level according to the rule of connectivity corresponding to the connecting regions, their granularity index and their laminar composition (as shown in table 2).
2. Assumed connections: vertically oriented connections between two local laminar components were modelled according to the laminar composition of each cortical region (as shown in figure 5).

It is worth noting that tractography connections represent the majority of connections in our model and they are also considerably stronger than the assumed connections.

The resulting network of cortical laminar connectivity has three times the nodes of the original network, since every regional node now includes three laminar locations in that same specific region. Additionally, each laminar group is now represented in a different location in the vertical axis. In order to better visualize the resulting laminar-level connectome, the connections in our model were colored according to the connecting laminar groups, including six categories such as infragranular-infragranular, infragranular-granular, etc. (see figure 8). Using an expanded or multilayered circular network graphs enables us to better examine and visualize both our model input and its output.

**Fig. 8.**
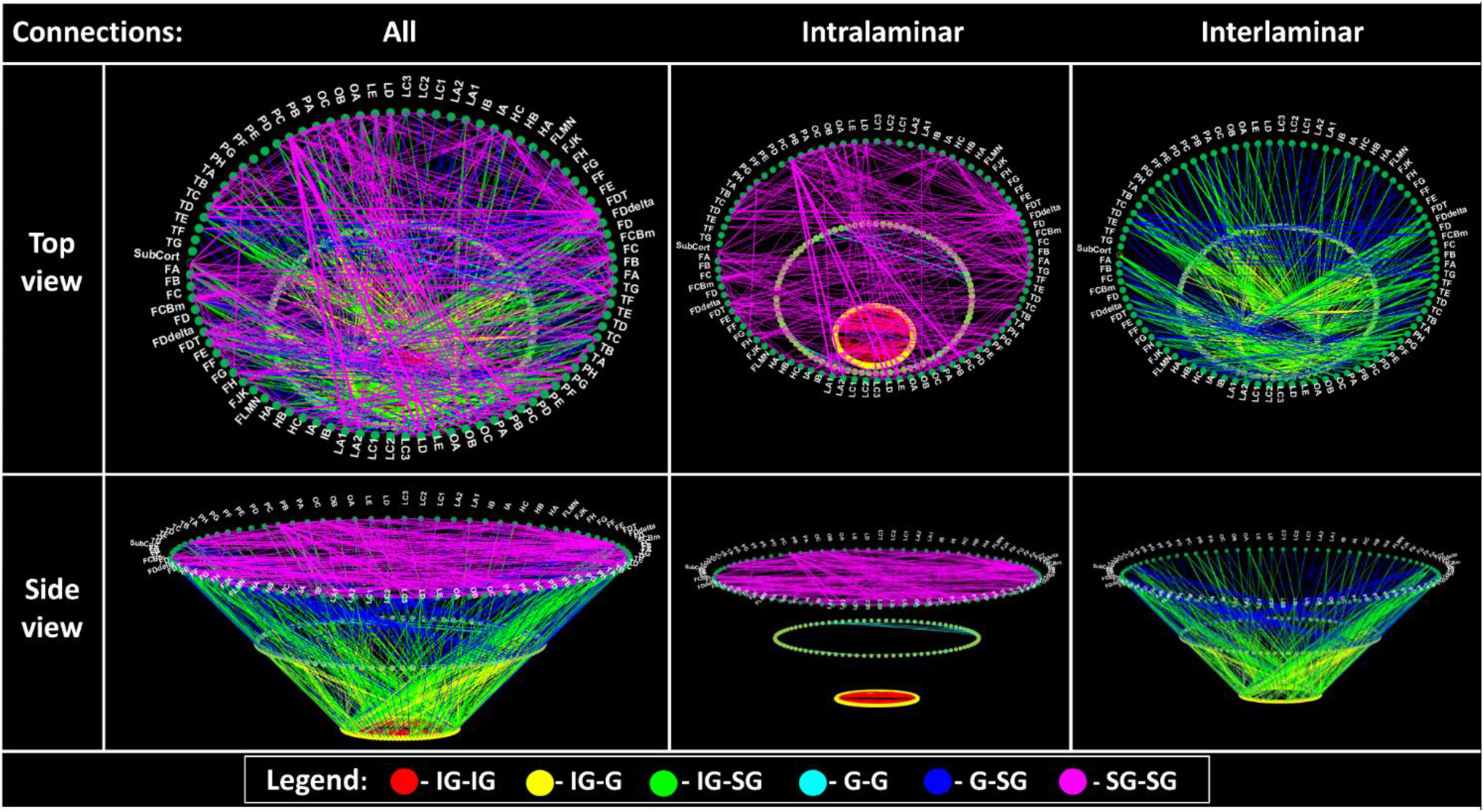
A multilayered circular network graph representation of our resulting model of cortical laminar connectivity. Columns: first column represents all model connections, second represents connections within a laminar group, and the third represents connections between laminar groups; Rows: top row includes a top view of the network graph, while the bottom row includes a side view; Legend: connections were colored according to connection type, based on the connecting laminar groups (visualization completed based on: Paul Kassebaum (2020), circularGraph (https://www.github.com/paul-kassebaum-mathworks/circularGraph), GitHub)

In short, our study offers a novel whole-brain approach to integrating white matter connectivity (DWI tractography) with cortical grey matter structure (T1 cortical laminar analysis), by incorporating measured macroscale connections with presumed microscale connections.

## 4 Discussion

Over the past decades, exploration of the cerebral cortex and its connectivity patterns has become an ever-growing focus of interest and research in the field of neuroscience. Exploration of the cortex includes study of its laminar structure, while exploration of cortical connectivity includes both interregional connectivity as well as intraregional laminar connectivity. This review aims to address the lingering ambiguity around the discussion of laminar representations and models of laminar interconnectivity, focusing on the various ways of partitioning the cortex into layers and defining its vertical and horizontal connections. Our emphasis is cortical laminar connectivity on both the morphology level as well as the connectivity level, as reported in 51 prominent published studies and reviews on the subject, primarily since the early 2000s.

Our model is based on a review of a limited number of articles, particularly compared to wider-scale integrative reviews of hundreds of articles on the topic primate laminar connectivity in specific cortical regions (Solari and Stoner 2011, Schmidt et al. 2018). While such reviews offer a broader and more in-depth viewpoint on the topic, our review is uniquely designed with the novel purpose of forming a model that integrate MRI datasets and overcomes their current limitations.

While our model application is conducted on human cortical data, it is in fact based on data from both human and nonhuman models. The reason for this circumstance is twofold: firstly, most of the published data on the subject uses a variety of methods that can only be applied on nonhuman animal models, and secondly is the current resolution limitation in in vivo imaging of cortical laminar connectivity on a microscale level. Nevertheless, the reviewed articles showcase the complexity of the subject of cortical layer connectivity, stemming from the. A wide variety of methods, definitions and outlooks is applied when addressing the subject of cortical laminar connectivity. The methods used for examining cortical layer connectivity are applied on both human and varying animal models, mainly macaque and cat (for a comparative review of neocortical architecture among regions and species see DeFelipe et al. 2002), and include histological staining, retrograde and anterograde tracers, electrical recording, MRI imaging and many more.

The relevant definitions even vary regarding Brodmann’s “classic” 1909 definition of six cortical layers, which are more highly differentiated in granular cortical regions. Cortical layers are grouped in different ways: with rough grouping into three groups up to a finer grouping into eight groups. Definitions of the layer connections also vary, examining a wide spectrum of connections, including input and output, feedback and feedforward, interlaminar and intralaminar connections and many more. With regards to the outlooks applied to the subject of cortical layer connectivity, the subject is typically approached on a local or region-specific level, focusing on a limited cortical region instead of a whole-brain level.

Our systematic review offers a simplified way to overcome some of these limitations by standardizing the reviewing process of reported laminar connections and their analysis. We unify and generalize our approach by adopting a granularity-based approach to both cortical layer grouping into infragranular, granular and supragranular layers, as well as cortical atlas selection, using the von Economo granularity atlas. By doing so, we can estimate connection probabilities of all reported cortical laminar connections. We offer a data-derived way to quantitatively and qualitatively model cortical laminar connectivity in a unified whole-brain manner.

Nowadays, exploration of the brain’s connectivity (also known as connetomics) has become a vast subject of research in the neuroimaging community. Despite the growing interest in connectomics, an inherent bias still exists in the field, due to the definition of the cortex as one homogeneous unit. Overcoming this bias demands expanding the basic unit used for connectome analysis to a more descriptive representation of the heterogeneous laminar substructure of the cortex.

So far, neuroimaging research has explored three main types of brain connectivity:

a. Structural connectivity: represents the anatomical links, or fiber bundles, connecting different cortical regions. Structural connectivity can be measured using DWI tractography (other non-MRI based techniques include retrograde tracing, such as the large-scale macaque study conducted by Ercsey-Ravasz et al. 2013).
b. Functional connectivity: represents time series correlation between different cortical regions. Functional connectivity can be measured using BOLD fMRI.
c. Effective connectivity: represents the influence one cortical region exerts over another. It is based on a network model of causal dynamics that best explains the functional signals (state-dependent), while being constrained by the anatomy of structural connectivity (Friston et al. 2003, Friston et al. 2019).

Cortical laminar connectivity represents a possible fourth type of brain connectivity that takes into account the heterogenous laminar structure of the cortex. It could be considered a model expansion of whole-brain structural connectivity, similarly to the way in which effective connectivity is a model expansion of functional connectivity. The division into laminar-level connectivity can help in refining observations and results from global tractography connectomics using graph indices. For example, rich-club organization and connectivity hubs (van den Heuvel and Sporns 2011, 2013) can be further explored using the laminar connectivity distinction and help shed more light on why the brain organizes this way.

We propose a unified whole-brain method to model cortical layer connectivity that is applicable to the limitations of MRI methodologies. Our model offers a way to integrate two neuroimaging datasets: global white matter connectivity (tractography) extracted from diffusion weighted imaging, and cortical grey matter structure (cortical laminar analysis) extracted from T1 imaging. The model incorporates macroscale white matter connections from tractography with presumed microscale connections on the cortical layer level, based on measured cortical laminar composition.

Our model utilizes the granularity-based approach to cortical layer grouping into infragranular, granular and supragranular layers, offering a robust laminar categorization of the cortex. While the model does not offer individual region-specific models of cortical circuitry, it offers a more generalized whole-brain microcircuit model that is adapted regionally based on an atlas of granularity indices. In other words, the model is both simplified and generalized using assumptions regarding circuity in differently granulated brain regions.

Standardization of the approach towards both cortical layers as well as cortical columns enabled the formation of our whole-brain model of cortical layer connectivity, which in turn enables us to bridge the gap between two branches of neuroimaging and offers a uniform way to approach and model mesoscale-level connectivity. Use of our offered framework could potentially expand the current study of connectomics and introduce new ways to model multilayered (or multidimensional) networks of brain connectivity.

Though this study may not include a neuroscientific breakthrough result, what it does offer is a novel modelling concept and an accompanying detailed framework for applying it. Our proposed model demonstrates that MRI limitations can be overcome to explore connectomics on the laminar level. Follow-up region-specific studies, focusing for example on the motor or visual cortices, could propose neuroscientific validation by comparison to published results, such as Felleman and Van Essen’s extensive work on the hierarchical structure of the visual cortex (Felleman and Van Essen 1991). Each such validation study could apply our framework with certain adaptations to the model.

## Data Availability

The source code of the present model connectomes (standard and laminar) and the codes for the circular graphs (expanded and multilayered, respectively) will be freely available for non-commercial use from GitHub.

## References

1. Hagmann P., Cammoun L., Gigandet X., Meuli R., Honey C. J., Wedeen V. J., Sporns O. (2008). Mapping the structural core of human cerebral cortex, PLoS biology 6(7), 1479–1493. doi:10.1371/journal.pbio.0060159.g001

2. Sporns O. (2012). Discovering the Human Connectome, MIT press, ISBN: 978-0-262-01790-9.

3. von Economo C. (2009). Cellular Structure of the Human Cerebral Cortex, ed L. C. Triarhou Karger Medical and Scientific Publishers, Basel. doi: 10.1159/000226273

4. Leemans A., Jeurissen B., Sijbers J., Jones D. K. (2009). ExploreDTI: a graphical toolbox for processing, analyzing, and visualizing diffusion MR data, In: 17th Annual Meeting of International Society of Magnetic Resonance in Medicine, p. 3537, Hawaii, USA.

5. Sporns O., Tononi G., Kotter R. (2005). The Human Connectome: A Structural Description of the Human Brain, PLoS Computational Biology 1(4): e42, 0245–0251. doi: 10.1371/journal.pcbi.0010042

6. Paolo G. P., Verma R., Lee S. K., Melhem E. R. (2007). Diffusion-Tensor MR Imaging and Tractography: Exploring Brain Microstructure and Connectivity, Radiology, Vol. 245, Num. 2, 367–384.

7. Van Essen D. C., Smith S. M., Barch D. M., Behrens T. E. J., Yacoub E., Ugurbil K.. (2013). The WU-Minn Human Connectome Project: An Overview, NeuroImage, 80, 62–79. doi: 10.1016/j.neuroimage.2013.05.041

8. Setsompop K., Kimmlingen R., Eberlein E., Witzel T., Cohen-Aded J., McNab J. A., Keil B., Tisdall M. D., Hoecht P., Dietz P., Cauley S.F., Tountcheva V., Matschl V., Lenz V. H., Heberlein K., Potthast A., Thein H., Van Horn J., Toga A., Schmitt F., Lehne D., Rosen B. R., Wedeen V., Wald L. L. (2013). Pushing the limits of in vivo diffusion MRIfor the Human Connectome Project, Neurolmage, 80. 220–233. doi: 10.1016/j.neuroimage.2013.05.078

9. Sotiropoulos S. N., Zalesky A. (2019). Building connectomes using diffusion MRI: why, how and but, NMR in Biomedicine, Vol. 32, iss. 4, 1–23. doi: 10.1002/nbm.3752

10. Clark V. P., Courchesne E., Grafe M. (1992). In vivo myeloarchitectonic analysis of human striate and extrastriate cortex using magnetic resonance imaging, Cerebral Cortex, 2: 417–424. doi: 10.1093/cercor/2.5.417

11. Van Essen D. C., Donahue C. J., Coalson T. S., Kennedy H., Hayashi T., Glasser M. F. (2019). Cerebral cortical folding, parcellation, and connectivity in humans, nonhuman primates, and mice, PNAS, vol. 116, no. 52, 26173-26180. doi: 10.1073/pnas.1902299116

12. Barazany D., Assaf Y. (2012). Visualization of Cortical Lamination Patterns with Magnetic Resonance Imaging, Cerebral Cortex, 22, 2016–2023. doi: 10.1093/cercor/bhr277

13. Lifshits S., Tomer O., Shamir I., Barazany D., Tsarfaty G., Rosset S., Assaf Y. (2018). Resolution considerations in imaging of the cortical layers, Neuroimage, 164, 112–120. doi: 10.1016/j.neuroimage.2017.02.086

14. Shamir I., Tomer O., Baratz Z., Tsarfaty G., Faraggi M., Horowitz A., Assaf Y. (2019). A framework for cortical laminar composition analysis using low-resolution T1 MRI images, Brain Structure and Function, vol. 224, is. 4, 1457–1467. doi: 10.1007/s00429-019-01848-2

15. Jbabdi S., Johansen-Berg J. (2011). Tractography: Where Do We Go from Here?, Brain Connectivity, Vol. 1, Num. 3, 169-183. doi: 10.1089/brain.2011.0033

16. Johansen-Berg H. (2013). Human connectomics – What will the future demand?, NeuroImage, 80, 541–544. doi: 10.1016/j.neuroimage.2013.05.082

17. Gennari F. (1782). De Peculiari Structura Cerebri, nonulisque ejus mrobis. Parma: Ex Regio Typographeo.

18. Baillarger J. G. F. (1840). Recherches sur la structure de la couche corticale des circonvolutions du cerveau, J. B. Bailliere.

19. Garey L. J. (2006). Brodmann’s localization in the cerebral cortex, Springer: Berlin 1–58. doi: 10.1142/p151

20. Lorente de Nó R. (1949). Cerebral cortex: architecture, intracortical connections, motor projections, Physiology of the Nervous System, ed Fulton J. F., editor. (New York, NY: Oxford University Press), 288–312.

21. Mountcastle V. B. (1957). Modality and topographic properties of single neurons of cat’s somatic sensory cortex, Journal of Neurophysiology. 20 (4): 408–34. doi: 10.1152/jn.1957.20.4.408. PMID 13439410.

22. Hubel D. H., Wiesel T. N. (1959). Receptive fields of single neurones in the cat’s striate cortex, Journal of Physiology, 148: 574–591.

23. Hubel D. H., Wiesel T. N. (1962). Receptive fields, binocular interaction and functional architecture in the cat’s visual cortex, Journal of Physiology, 160: 106–154.

24. Hubel D. H., Wiesel T. N. (1965). Receptive fields and functional architecture in two nonstriate visual areas (18 and 19) of the cat, Journal of Neurophysiology, 28: 229–289.

25. Hubel D. H., Wiesel T. N. (1968). Receptive fields and functional architecture of monkey striate cortex, Journal of Physiology, 195: 215–243.

26. Hubel D. H., Wiesel T. N. (1969). Visual area of the lateral suprasylvian gyrus (Clare-Bishop area) of the cat, Journal of Physiology, 202: 251–260.

27. Szentágothai J. (1975). The ‘module-concept’ in cerebral cortex architecture, Brain Research, 95: 475–496.

28. Hellweg F. C., Schultz W., Creutzfeldt O. D. (1977). Extracellular and intracellular recordings from cat’s cortical whisker projection area: thalamocortical response transformation, Journal of Neurophysiology, 40(3): 463–479.

29. Gilbert C. D., Wiesel T. N. (1983). Functional organization of the visual cortex, Progress in Brain Research 58: 209–18.

30. Rockland K. S., Pandya D. N. (1979). Laminar origins and terminations of cortical connections of the occipital lobe in the rhesus monkey, Brain Research: 179(1): 3–20. doi: 10.1016/0006-8993(79)90485-2

31. Maunsell J. H., Van Essen D. C. (1983). Functional properties of neurons in middle temporal visual area of the macaque monkey. I. Selectivity for stimulus direction, speed, and orientation, Journal of Neurophysiology 49(5): 1127–47. doi: 10.1152/jn.1983.49.5.1127

32. Felleman D. J., Van Essen D. C (1991). Distributed Hierarchical Processing in the Primate Cerebral Cortex, Cerebral Cortex, 1, 47–1.

33. Solari S. V. H., Stoner R. (2011). Cognitive consilience: primate non-primary neuroanatomical circuits underlying cognition, Frontiers in Neuroanatomy, vol. 4, article 65, 1–23. doi: 10.3389/fnana.2011.00065

34. Schmidt M., Bakker R., Hilgetag C. C., Diesmann M., Van Albada S. J. (2018). Multi-scale account of the network structure of macaque visual cortex, Brain Structure and Function, 223: 1,409–1,435. doi: 10.1007/s00429-017-1554-4

35. Douglas R. J., Martin K. A. C., Whitteridge D. (1989). A Canonical Microcircuit for Neocortex, Neural Computation, 1, 480–488.

36. Moher D., Liberati A., Tetzlaff J., Altman D. G., The PRISMA Group (2009). Preferred Reporting Items for Systematic Reviews and Meta-Analyses: The PRISMA Statement, PLoS Medicine 6 (7): e1000097. doi: 10.1371/journal.pmed1000097

37. Krzywinski M. I., Schein J. E., Birol I., Connors J., Gascoyne R., Horsman D., Jones S. J., Marra M. A. (2009). Circos: An information aesthetic for comparative genomics, Genome Research, Published in Advance June 18, 2009. doi:10.1101/gr.092759.109

38. Scholtens L. H., de Reus M. A., de Lange S. C., Schmidt R., van den Heuvel M. P. (2016). An MRI Von Economo-Koskinas atlas, NeuroImage, 170: 249–256. doi: 10.1016/j.neuroimage.2016.12.069

39. Friston K. J., Harrison L., Penny W. (2003). Dynamic causal modelling, NeuroImage, vol. 19(4): 1273–1302. doi: 10.1016/S1053-8119(03)00202-7

40. Friston K. J., Preller K. H., Mathys C., Cagnan H., Heinzle J., Razi A., Zeidman P. (2019). Dynamic causal modelling revisited, NeuroImage 199: 730–744. doi: 10.1016/j.neuroimage.2017.02.045

41. Ercsey-Ravasz M., Markov N. T., Lamy C., Van Essen D. C., Knoblauch K., Toroczkai Z., Kennedy H. (2013). A Predictive Network Model of Cerebral Cortical Connectivity Based on a Distance Rule, Neuron, 80: 184–197. doi: 10.1016/j.neuron.2013.07.036

42. Barone P., Batardiere A., Knoblauch K., Kennedy H. (2000). Laminar Distribution of Neurons in Extrastriate Areas Projecting to Visual Areas V1 and V4 Correlates with the Hierarchical Rank and Indicates the Operation of a Distance Rule, The Journal of Neuroscience, 20(9):3263–3281. doi: 10.1523/JNEUROSCI.20-09-03263.2000

43. Donahue C. J., Sotiropoulos S. N., Jbabdi S., Hernandez-Fernandez M., Behrens T. E., Dyrby T. B., Coalson T., Kennedy H., Knoblauch K., Van Essen D. C., Glasser M. F. (2016). Using Diffusion Tractography to Predict Cortical Connection Strength and Distance: A Quantitative Comparison with Tracers in the Monkey, The Journal of Neuroscience, 36(25): 6758–6770. doi: 10.1523/JNEUROSCI.0493-16.2016

44. Van den Heuvel M. P., Sporns O. (2011). Rich-club Organization of the Human Connectome, Journal of Neuroscience, 31(44), 15775–15786. doi: 10.1523/JNEUROSCI.3539-11.2011.

45. Van den Heuvel M. P., Sporns O. (2013). Network hubs in the human brain, Trends in Cognitive Sciences, 17(12), 683:696. doi: 10.1016/j.tics.2013.09.012

## Included in systematic review

46. Roelfsema P. R., Holtmaat A. (2018). Control of synaptic plasticity in deep cortical networks, Nature Reviews Neuroscience, 19, 180–166. doi: 10.1038/nrn.2018.6

47. Felleman D. J., Van Essen D. C (1991). Distributed Hierarchical Processing in the Primate Cerebral Cortex, Cerebral Cortex, 1, 47–1.

48. Da Costa N. M., Martin K. A. C. (2010). Whose cortical column would that be?, Frontiers in Neuroanatomy, Vol. 4, Article 16, 10–1. doi: 0.3389/fnana.2010.00016

49. Binzegger T., Douglas R. J., Martin K. A. C. (2004). A Quantitative Map of the Circuit of Cat Primary Visual Cortex, The Journal of Neuroscience, (39) 24, 8453–8441. doi: 10.1523/JNEUROSCI.1400-04.2004

50. Douglas R. J., Martin K. A. C. (2007). Mapping the Matrix: The Ways of the Neocortex, Neuron, 56, 238–226. doi: 10.1016/j.neuron.2007.10.017

51. Dhruv N. T. (2015). Rethinking canonical cortical circuits, Nature Neuroscience, 18, 1538. doi: 10.1038/nn1115-1538

52. Nelson S. B. (2002). Cortical Microcircuits: Diverse or Canonical?, Neuron, 36, 27–19. doi: 10.1016/S0896-6273(02)00944-3

53. Beul S. F., Hilgetag C. C. (2015). Towards a “canonical” agranular cortical microcircuit, Frontiers in Neuroanatomy, 165, 8. doi: 10.3389/fnana.2014.00165

54. Douglas R. J., Martin K. A. C. (1991). A Functional Microcircuit for Cat Visual Cortex, Journal of Physiology, 440, 769–735.

55. Hubel D. H., Wiesel T. N. (1962). Receptive Fields, Binocular Interaction and Functional Architecture in the Cat’s Visual Cortex, Journal of Physiology, 160, 154–106.

56. Raizada R. D. S., Grossberg S. (2003). Towards a Theory of the Laminar Architecture of Cerebral Cortex: Computational Clues from the Visual System, Cerebral Cortex, Volume 13, Issue 1, 113–100. doi: 10.1093/cercor/13.1.100

57. Thomson A. M. (2010). Neocortical layer 6, a review, Frontiers in Neuroanatomy, Volume 4, Article 13, 83–70. doi: 10.3389/fnana.2010.00013

58. Kiernan J., Rajakumar N. (2014). Barr’s The Human Nervous System - An Anatomical Viewpoint, Lippincott Williams & Wilkins, 246–213, 62–13.

59. Estrada-Sanchez A., Rebec G. V. (2013). Role of cerebral cortex in neuropathology of Huntington’s disease, Frontiers in Neural Circuits, Volume 7, Article 19, 9–1. doi: 10.3389/fncir.2013.00019

60. Guy J., Staiger J. F. (2017). The Functioning of a Cortex without Layers, Frontiers in Neuroanatomy, Volume 11, Article 54, 13–1. doi: 10.3389/fnana.2017.00054

61. Lubke J., Feldmeyer D. (2007). Excitatory signal flow and connectivity in a cortical column: focus on barrel cortex, Brain Structure and Function, 212, 17–3. doi: 10.1007/s00429-007-0144-2

62. Mumford D. (1991). On the computational architrcutre of the neocortex, Biological Cybernetics, (2) 65, 145–135. doi: 10.1007/BF00202389

63. Mountcastle V. B. (1997). The columnar organization of the neocortex, Brain, 120, 722–701.

64. Fishell G., Rudy B. (2011). Mechanisms of Inhibtion within the Telecephalon: “Where the Wild Things Are”, Annual Review of Neuroscience, 34, 567–535. doi: 10.1146/annurev-neuro-061010-113717

65. Mercer A., Thomson A. M. (2017). Cornu Ammonis Regions-Antecedents of Cortical Layers?, Frontiers in Neuroanatomy, Volume 11, Article 83, 25–1 doi: 10.3389/fnana.2017.00083

66. Helmstaedter M., Staiger J. F., Sakmann B., Feldmeyer D. (2008). Efficient Recruitment of Layer 2/3 Interneurons by Layer 4 Input in Single Columns of Rat Somatosensory Cortex, The Journal of Neuroscience, (33) 28, 8284–8273. doi: 10.1523/JNEUROSCI.5701-07.2008

67. Shepherd G. M. (2011). The microcircuit concept applied to coritcal evolution: from three-layer to six-layer cortex, Frontiers in Neuroanatomy, Volume 5, Article 30, 15–1. doi: 10.3389/fnana.2011.00030

68. Douglas R. J., Martin K. A. C. (2004). Neuronal Circuits of the Neocortex, Annual Review of Neuroscience, 27, 419–451. doi: 10.1146/annurev.neuro.27.070203.144152

69. Binzegger T., Douglas R. J., Martin K. A. C. (2009). Topology and dynamics of the canonical circuit of cat V1, Neural Networks, 22, 1078–1071. doi: 10.1016/j.neunet.2009.07.011

70. Helmstaedter M., de Kock C. P. J., Feldmeyer D., Bruno R. M., Sakmann B. (2007). Reconstruction of an average cortical column in silico, Brain Research Reviews, 55, 203–193. doi: 10.1016/j.brainresrev.2007.07.011

71. Feldmeyer D. 2012. Excitatory neuronal connectivity in the barrel cortex, Frontiers in Neuroanatomy Volume 6, Article 25, 22–1. doi: 10.3389/fnana.2012.00024

72. Harris K. D., Mrsic-Flogel T. D. (2013). Cortical connectivity and sensory coding, Nature, 503, 58–51. doi:10.1038/nature12654

73. Thomson A. M., Lamy C. (2007). Functional Maps of Neocortical Local Circuitry, Frontiers in Neuroscience, Volume 1, Issue 1, 42–19. doi: 10.3389/neuro.01.1.1.002.2007

74. Shipp S. (2007). Structure and function of the cerebral cortex, Current Biology, Volume 17, Issue 12, 449–443. doi: 10.1016/j.cub.2007.03.044

75. Herkenham M. (1980). Laminar Organization of Thalamic Projections, Science, Volume 207, Issue 4430, 535–532. doi: 10.1126/science.7352263

76. Buxhoeveden D. P., Casanova M. F. (2002). The minicolumn hypothesis in neuroscience, Brain, Volume 125, Issue 5, 951–935. doi: 10.1093/brain/awf110

77. Lodato S., Arlotta P. (2015). Generating Neuronal Diversity in the Mammalian Cerebral Cortex, Annual Review of Developmental Biology, Volume 31, 720–699. doi: 10.1146/annurev-cellbio-100814-125353

78. Hawkins J., Ahmad S., Cui Y. (2017). A Theory of How Columns in the Neocortex Enable Learning the Structure of the World, Frontiers in Neural Circuits, Volume 11, Article 81, 18–1. doi: 10.3389/fncir.2017.00081

79. Du J., Vegh V., Reutens D. (2012). The Laminar Cortex Model: A New Continuum Cortex Model Incorporating Laminar Architecture, PLoS Computational Biology, (10) 8, 9–1. doi: 10.1371/journal.pcbi.1002733

80. Weiler N., Wood L., Yu J., Solla S. A., Shpherd G. M. G. (2008). Top-down laminar organization of the excitatory network in motor cortex, Nature, Neuroscience, 11, 366–360. doi:10.1038/nn2049

81. Shipp S. (2005). The importance of being agranular: a comparative account of visual and motor cortex, Philosophical transactions of the royal society, 360, 814–797. doi:10.1098/rstb.2005.1630

82. Bosman C. A., Aboitiz F. (2015). Functional constraints in the evolution of brain circuits, Frontiers in Neuroscience, Volume 9, Article 303, 13–1. doi: 10.3389/fnins.2015.00303

83. Rockland K. S. (2017). What do we know about laminar connectivity?, NeuroImage, xxx, 13–1. doi: 10.1016/j.neuroimage.2017.07.032

84. Douglas R. J., Martin K. A. C., Whitteridge D. (1989). A Canonical Microcircuit for Neocortex, Neural Computation, 1, 480–488.

85. Larkum M. E., Petro L. S., Sachdev R. N. S., Muckli L. (2018). A Perspective on Cortical Layering and Layer-Spanning Neuronal Elements, Frontiers in Neuroanatomy, Volume 12, Article 56, 1–9. doi: 10.3389/fnana.2018.00056

86. Bastos A. M., Usrey W. M., Adams R. A., Mangun G. R., Fries P., Friston K. J. (2012). Canonical microcircuits for predictive coding, Neuron, 76(4): 695–711. doi: 10.1016/j.neuron.2012.10.038

87. Meyer H. S., Wimmer V. C., Hemberger M., Bruno R. M., de Kock C. P. J., Frick A., Sakmann B., Helmstaedter M. (2010). Cell Type--Specific Thalamic Innervation in a Column of Rat Vibrissal Cortex, Cerebral Cortex, 20: 2287–2303. doi: 10.1093/cercor/bhq069

88. DeFelipe J., Alonso-Nanclares L., Arellano J. I. (2002). Microstructure of the neocortex: Comparative aspects, Journal of Neurocytology, 31: 299–316. doi: 10.1023/A:1024130211265

89. Meyer H. S., Wimmer V. C., Oberlaender M., de Kock C. P. J., Sakmann B., Helmstaedter M. (2010). Number and Laminar Distribution of Neurons in a Thalamocortical Projection Column of Rat Vibrissal Cortex, Cerebral Cortex, 20: 2277–2286. doi: 10.1093/cercor/bhq067

90. Jiang X., Shen S., Cadwell C. R., Berens P., Sinz F., Ecker A. S., Patel S., Tolias A. S. (2015). Principles of connectivity among morphologically defined cell types in adult neocortex, Science, 350(6264). doi: 10.1126/science.aac9462

91. Markram H., Muller E., Ramaswamy S., Reimann M. W., Abdellah M., Sanchez C. A., Ailamaki A., Alonso-Nanclares L., Antille N., Arsever S., Kahou G. A. A., Berger T. K., Bilgili A., Buncic N., Chalimourda A., Chindemi G., Courcol J. D., Delalondre F., Delattre V., Druckmann S., Dumusc R., Dynes J., Eilemann S., Gal E., Gevaert M. E., Ghobril J. P., Gidon A., Graham J. W., Gupta A., Haenel V., Hay E., Heinis T., Hernando J. B., Hines M., Kanari L., Keller D., Kenyon J., Khazen G., Kim Y., King J. G., Kisvarday Z., Kumbhar P., Lasserre S., Le Be’ J. V., Magalha B. R. C., Merchan-Perez A., Meystre J., Morrice B. R., Muller J., Munoz-Cespedes A., Muralidhar S., Muthurasa K., Nachbaur D., Newton T. H., Nolte M., Ovcharenko A., Palacios J., Pastor L., Perin R., Ranjan R., Riachi I., Rodriguez J. R., Riquelme J. L., Rossert C., Sfyrakis K. Shi Y., Shillcock J. C., Silberberg G., Silva R., Tauheed F., Telefont M., Toledo-Rodriguez M., Trankler T., Van Geit W., Diaz J. V., Walker R., Wang Y., Zaninetta S. M., DeFelipe J., Hill S. L., Segev I., Schurmann F. (2015). Reconstruction and Simulation of Neocortical Microcircuitry, Cell, 163: 456–492. doi: 10.1016/j.cell.2015.09.029

92. Ramaswamy S., Courcol J. D., Abdellah M., Adaszewski S. R., Antille N., Arsever S., Atenekeng G., Bilgili A., Brukau Y., Chalimourda A., Chindemi G., Delalondre F., Dumusc R., Eilemann S., Gevaert M. E., Gleeson P., Graham J. W., Hernando J. B., Kanari L., Katkov Y., Keller D., King J. G., Ranjan R., Reimann M. W., Rössert C., Shi Y., Shillcock J. C., Telefont M., Van Geit W., Diaz J. V., Walker R., Wang Y., Zaninetta S. M., DeFelipe J., Hill S. L., Muller J., Segev I., Schü;rmann F., Muller E. B., Markram H. (2015). The neocortical microcircuit collaboration portal: a resource for rat somatosensory cortex, Frontiers in Neural Circuits, vol. 9, article 44. doi: 10.3389/fncir.2015.00044

93. Markov N. T., Vezoli J., Chameau P., Falchier A. Quilodran R., Huissoud C., Lamy C., Misery P., Giroud P., Ullman S., Barone P., Dehay C., Knoblauch K., Kennedy H. (2014). Anatomy of Hierarchy: Feedforward and Feedback Pathways in Macaque Visual Cortex, The Journal of Comparative Neurology, Research in Systems Neuroscience, 522: 225–259. doi: 10.1002/cne.23458

94. Wang X. J., Kennedy H. (2016). Brain structure and dynamics across scales: In search of rules, Current Opinion in Neurobiology, 37: 92–98. doi:10.1016/j.conb.2015.12.010

95. Hirsch J. A., Martinez L. M. (2006). Laminar processing in the visual cortical column, Current Opinion in Neurobiology, 16: 377–384. doi: 10.1016/j.conb.2006.06.014

96. Kennedy H., Dehay C. (2012). Self-organization and interareal networks in the primate cortex, Progress in Brain Research, Vol. 195, 341–360. doi: 10.1016/B978-0-444-53860-4.00016-7

